# Evaluation of the Peterborough Public Health COVID-19 Rapid Antigen Test Self-Report Tool: Implications for COVID-19 Surveillance

**DOI:** 10.1101/2022.10.28.22281659

**Authors:** Erin Smith, Carolyn Pigeau, Jamal Ahmadian-Yazdi, Mohamed Kharbouch, Jane Hoffmeyer, Thomas Piggott

## Abstract

**Background:** The ongoing COVID-19 pandemic has necessitated novel testing strategies, including the use of Rapid Antigen Tests (RATs). The widespread distribution of RATs to the public prompted Peterborough Public Health to launch a pilot RAT self-report tool to assess its utility in COVID-19 surveillance.

**Objective:** To investigate the utility of a RAT self-report tool through an analysis of the temporal association between RAT results, PCR test results, and wastewater levels of COVID-19.

**Methods:** We investigated the association between RAT results, PCR test results, and wastewater levels of COVID-19 using Pearson’s correlation coefficient. Percent positivity and count of positive tests for RATs and PCR tests were analyzed.

**Results:** PCR percent positivity and wastewater were weakly correlated (*r*=0.33 *p*=0.022), as were RAT percent positivity and wastewater (*r*=0.33 *p*=0.002). RAT percent positivity and PCR percent positivity were not significantly correlated (*r=*-0.035, *p*=0.75). Count of positive RAT tests and count of positive PCR tests were moderately correlated (*r*=0.59, *p*<0.001). Wastewater was not significantly correlated to count of positive RAT tests (*r*=0.019, *p=*0.864) or count of positive PCR tests (*r*=0.004, *p*=0.971).

**Conclusion:** Our results provide evidence in support of the use of RAT self-reporting as a low-cost simple adjunctive COVID-19 surveillance tool, and may suggest that its utility is greatest when considering an absolute count of positive RAT tests rather than percent positivity due to reporting bias towards positive tests. These results can help inform COVID-19 surveillance strategies of local Public Health Units and encourage the use of a RAT self-report tool.

## Background

### Topic background

The ongoing COVID-19 pandemic has warranted the implementation of novel testing strategies. The widespread distribution of self-administered rapid antigen tests (RATs), which can detect the viral proteins that cause COVID-19 in as little as 15 minutes (1), is one method employed by the government of Ontario to monitor the virus’ spread. In December 2021, the Ontario government announced the distribution of RATs for elementary and secondary school students and staff (2), followed by the expansion of the RAT rollout to pharmacies and grocery stores in February 2022 (3). This program continues until present with wide availability of publicly-funded RATs in addition to the ability for residents to purchase privately.

The announcement of limits to polymerase chain reaction (PCR) testing for only high-risk individuals in late 2021 posed a new challenge for accurately monitoring COVID-19 case counts (4). Wastewater surveillance has proven useful in addressing surveillance gaps (5-7), however, presents limitations in interpretation. Restrictions on PCR testing as well as the provincial distribution of RATs prompted Peterborough Public Health (PPH) to launch a pilot surveillance project asking local residents to voluntarily and confidentially their RAT results via an online self-report tool. The objective was to monitor an approximate percent positivity among those who reported their results to contribute to community COVID-19 surveillance.

### Research background and study objective

Given that PPH is the only public health unit globally that we are aware of that has launched a RAT self-report tool, an investigation into the utility of such a tool was identified as needed. The objective of this study is to investigate the correlation between RAT self-report results and other indicators of COVID-19, specifically PCR test results and wastewater levels of COVID-19. This is the first study we are aware of to investigate the utility of a RAT self-report tool, and the results will contribute robust evidence towards the role of RAT self-report results in COVID-19 surveillance.

## Methods

### Methods summary/overview

We investigated the temporal association between RAT results, PCR test results, and wastewater levels of COVID-19 using Pearson’s correlation coefficient. Both percent positivity as well as count of positive tests for RAT and PCR tests were analyzed. Wastewater levels of COVID-19 were measured in the region’s primary municipal wastewater source, the City of Peterborough, using a normalized average of viral genes N1 and N2 (nmN1N2).

PCR tests results were obtained from the Ontario Ministry of Health and Long-Term Care COVID-19 Testing Dashboard (8). Wastewater samples were collected and analyzed by Trent University in Peterborough. RAT results were obtained from the PPH online RAT self-report tool (9). PCR and RAT results from Peterborough City and County were included in the analysis while wastewater data from the City of Peterborough were included.

### Ethics Approval

This study did not require ethics approval as the data we analyzed is routinely collected by PPH for COVID-19 surveillance and is not reported on an individually identifiable basis.

### Analysis

The relationship between RAT results, PCR results, and wastewater levels of COVID-19 was analyzed using Pearson’s correlation. The data obtained for all three indicators were collected from December 17, 2021-April 30, 2022; dates with missing data were excluded. All data were analyzed using R (10). RAT, PCR, and wastewater data were screened for outliers and log-transformed to meet the assumption of a normal distribution for Pearson’s correlation.

## Results

### Descriptive results

From December 17, 2021 to April 30, 2022, 4571 self-report responses were recorded; 2138 responses were reported as positive.

**Figure 1:**
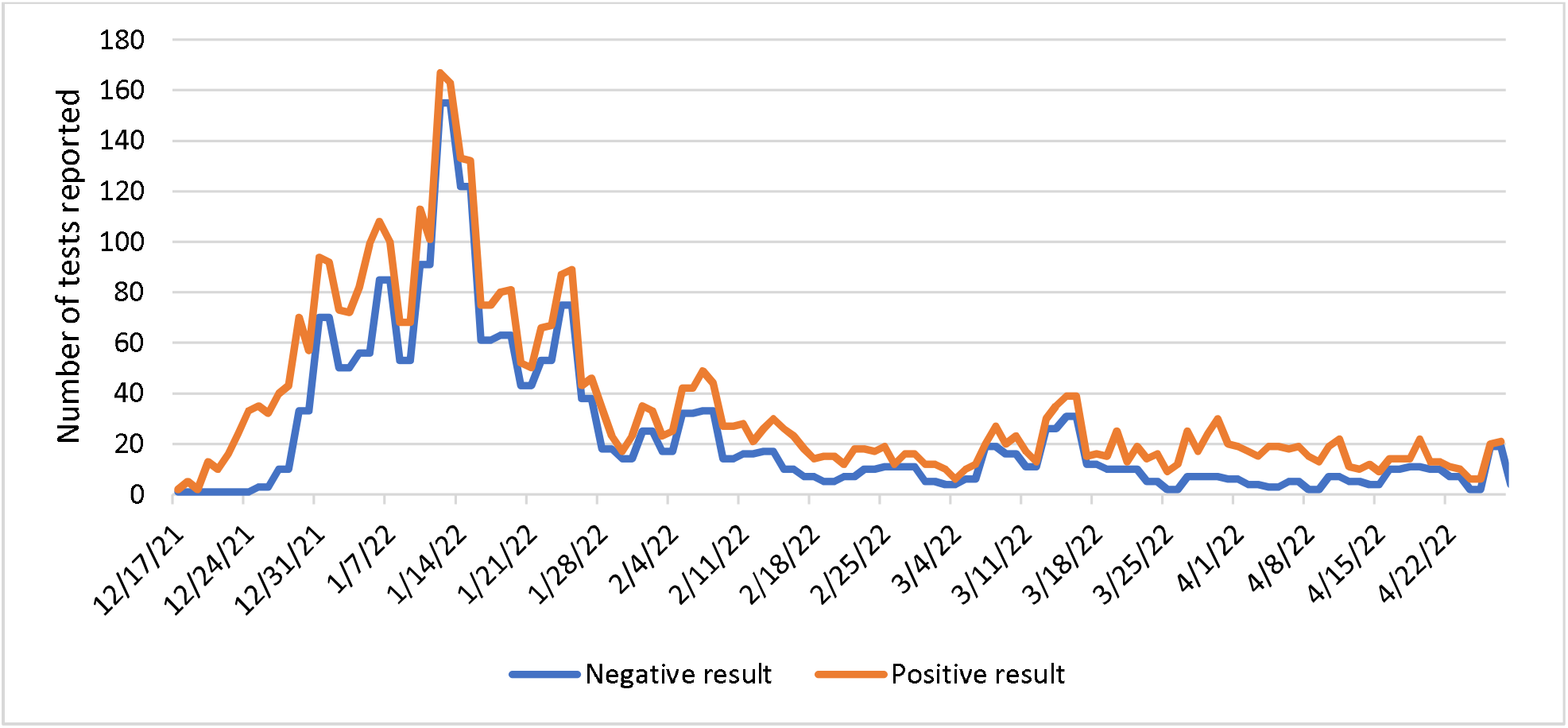
Count of reported positive and negative RAT results by date in the Peterborough Public Health region

**Figure 2:**
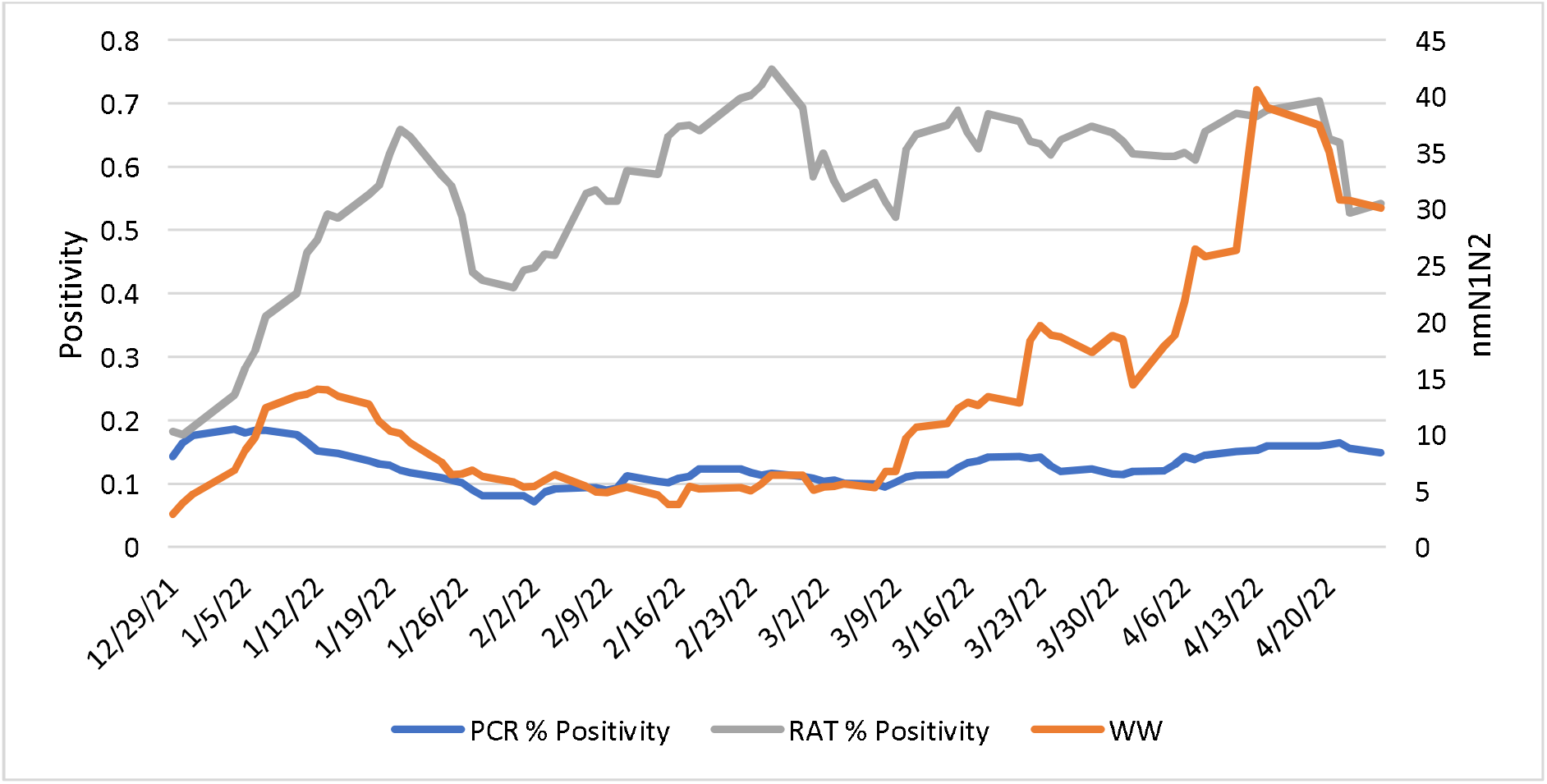
Percent Positivity of PCR Tests, Percent Positivity of RAT tests, and Normalized Viral Copies of COVID-19 by date in the Peterborough Public Health region

**Figure 3:**
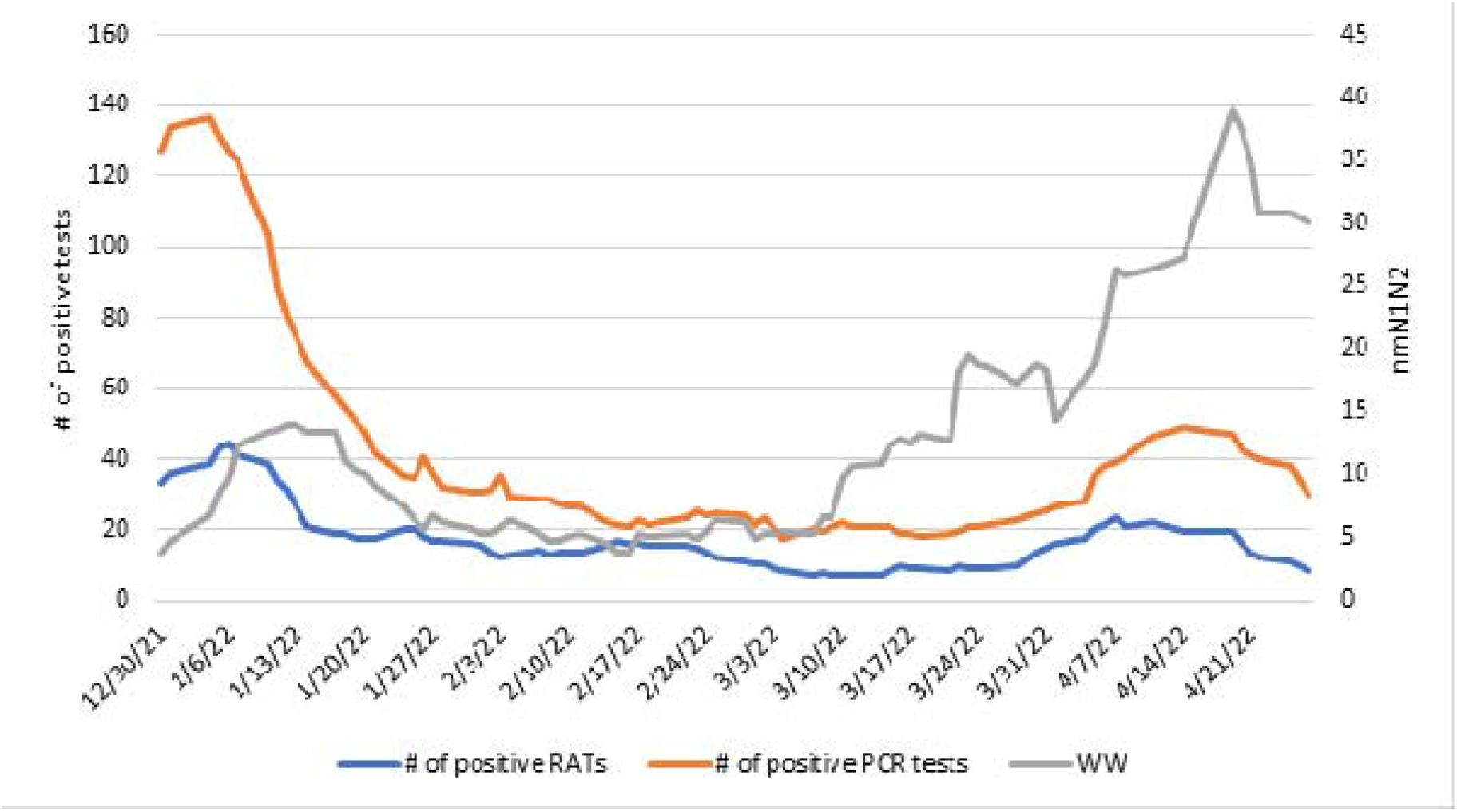
Count of Positive PCR Tests, Count of Positive RATs, and Normalized Viral Copies of COVID-19 in the Peterborough Public Health region

### Analytical results

When we analyzed the relationship between RAT percent positivity, PCR percent positivity, and wastewater levels, PCR percent positivity and wastewater were weakly correlated (*r*=0.33 *p*=0.022), as were RAT percent Positivity and wastewater (*r*=0.33 *p*=0.002); these results were statistically significant. RAT percent positivity and PCR percent positivity were not significantly correlated to each other (Table 1).

**Table 1:** Correlation Matrix of RAT Percent Positivity, PCR Percent Positivity, and Wastewater levels from December 17, 2021-April 30, 2022 (Pearson’s correlation, *r*).

|  | RAT % Positivity | PCR % Positivity | Wastewater (nmN1N2) |
| --- | --- | --- | --- |
| RAT % Positivity | 1.0 | -0.035($p=0.75$) | **0.33** ($p=0.002$) |
| PCR % Positivity | -0.035 ($p=0.75$) | 1.0 | **0.33** ($p=0.022$) |
| Wastewater (nmN1N2) | **0.33** ($p=0.002$) | **0.33** ($p=0.022$) | 1.0 |

When we analyzed the relationship between count of positive RAT tests, count of positive PCR tests, and wastewater levels, count of positive RAT tests and count of positive PCR tests were moderately correlated (*r*=0.59, *p*<0.001); this result was statistically significant. Wastewater was not significantly correlated to count of positive RAT tests or count of positive PCR tests (Table 2).

**Table 2:** Correlation Matrix of count of positive RAT, count of positive PCR, and Wastewater levels from December 17, 2021-April 30, 2022 (Pearson’s correlation, *r*).

|  | Count of positive RAT | Count of positive PCR | Wastewater (nmN1N2) |
| --- | --- | --- | --- |
| Count of positive RAT | 1.0 | **0.59** ($p<0.001$) | 0.019 ($p=0.864$) |
| Count of positive PCR | **0.59** (p<0.001) | 1.0 | 0.004 (p=0.971) |
| Wastewater (nmN1N2) | 0.019 (p=0.864) | 0.004 (p=0.971) | 1.0 |

## Discussion

### Overview

Our study is the first to investigate the utility of a RAT self-report tool for COVID-19 surveillance. We found that both PCR percent positivity and RAT percent positivity were weakly correlated to wastewater levels of COVID-19, and the count of positive RAT and count of positive PCR tests were moderately correlated. We did not find a significant correlation between PCR percent positivity and RAT percent positivity, nor the count of positive RATs and count of positive PCR tests with wastewater. A possible explanation for these results is that not everyone who contracts COVID-19 obtains a RAT or PCR testing any longer, many cases are asymptomatic or testing access criteria has changed. In addition, a significant proportion of people with active COVID-19 infections shed the virus in their stool, sometimes before their symptoms start; this contrasts with the time lag associated with RAT and PCR testing (11). Previous studies have also indicated the incidence and persistence of viral shedding through feces even after a negative nasopharyngeal swab (12). These temporal differences, along with a lack of testing in some active cases, may offer explanation for why there was no significant correlation observed between the counts of positive PCR or RAT tests with wastewater.

### Strengths

Our study is strong in that we used data collected during the peak in cases during the Omicron wave in Ontario. Heightened public awareness of COVID-19 transmission as well as the launch of the PPH RAT self-report tool contributed to a high number of RAT self-report submissions and an overall robust dataset. In a complex landscape of COVID-19 surveillance, RAT self-reporting may present a helpful adjunctive surveillance tool at a minimal cost and administrative burden to public health organizations.

### Limitations

The main limitation in our study is the bias in reporting COVID-19 cases through both PCR testing and the RAT self-report tool. As mentioned, the restrictions on PCR testing to only high-risk individuals in late 2021 likely resulted in a significant under-representation of actual COVID-19 cases in the PCR data that we analyzed. In addition, the voluntary nature of self-reporting a RAT result introduces inaccuracies, as the PPH self-report tool likely only captured a fraction of actual positive and negative RAT results. Intermittent efforts to raise awareness of RAT self-reporting, and advertising may result in fluctuations when the baseline reporting rates we observed were fairly low. A key limitation also relates to availability of RATs free-of-charge. At this stage provincial availability of RATs is set to continue until the end of 2022. If freely available tests cease, the likely respondents would be further restricted to those who could afford or choose to purchase RATs. The role of differential positive reporting bias in the RAT self-report data necessitated the analysis of absolute numbers of positive RAT tests and trends over time as opposed to traditional measures such as percent positivity.

Another limitation in our study is that the wastewater data we analyzed was collected only from the City of Peterborough; these samples may have differed from other areas in the region. However, the City of Peterborough represents the majority of the PPH population and can therefore be used as a proxy for the PPH region in a region geographically small such as ours.

### Implications for policy

The weak correlation that we found between RAT percent positivity and wastewater as well as PCR percent positivity and Wastewater suggests that RAT percent positivity can be used in COVID-19 surveillance; however, must be interpreted relative to limitations. Further assessment through additional waves of COVID-19 is needed to understand temporality in RAT self-reporting signals. For instance, corroborative information of RAT percent positivity, PCR percent positivity and increasing wastewater signals may be useful in reporting on changes in community transmission risk. Indeed, this is the reason that PPH has incorporated RAT testing into its COVID-19 Community Risk Index (13).

The moderate correlation between count of positive RAT and count of positive PCR tests suggests that the utility of RAT self-report results is greatest when using the count of positive RAT tests compared to percent positivity as a tool for COVID-19 surveillance due to reporting biases. In the event of further changes to PCR testing eligibility continuing to decrease, maintaining access to RAT and RAT self-reporting could be a complementary surveillance tool. These results could provide evidence in favour of the use of RAT self-report results in COVID-19 surveillance and support the potential for Public Health Units across Canada to develop local RAT self-report tools.

With this being said, multiple surveillance tools are likely required to obtain the most accurate picture of pandemic risk. For example, PPH has developed a COVID-19 Risk Index to inform community members about the current risk for COVID-19 transmission (13). Six indicators, one of which is a count of positive RATs, are weighted to present an overall risk level with accompanying risk guidance.

In addition, some limitations surrounding bias might be effectively addressed through collaboration between the public funder of RATs and local public health agencies (LPHA). For example, exploring the potential for linking RAT promotion, distribution and surveillance through a coordinated mechanism, such as including a QR code on the test package linking to a survey.

### Implications for research

Given that we identified reporting biases as a risk to the use of data from RAT self-reports, further research is required to determine how Public Health Units can encourage community members to report their RAT results. Additionally, further research on alternative surveillance tools is important as we progress through the pandemic given decreased access to PCR testing and reliability of their results to measure community transmission. Finally, additional research is warranted to investigate which indicators are the most sensitive to change. From a visual analysis of our data during wave 6 (March-April 2022), wastewater was the most sensitive as it was the first indicator to increase. RAT results have the next greatest sensitivity, with PCR being the least sensitive as it was the last indicator to show an increase. Further research evaluation is required to assess the sensitivity of each indicator to enable the earliest detection of an increase in COVID-19.

## Conclusions

The ongoing COVID-19 pandemic has necessitated the use of numerous indicators and testing strategies to approximate levels of COVID-19 transmission. Our results provide evidence in support of the use of RAT self-reports in COVID-19 surveillance. The utility of RAT self-reports is likely greatest when considering count of positive RAT tests compared to percent positivity due to reporting biases. The results from our study can help inform the COVID-19 surveillance strategies of local Public Health Units and encourage the use of a RAT self-report tool.

## Data Availability

All data produced in the present study are available upon request to the authors

## Author’s statement

ES- Analysis, writing, original draft, review, editing

CP- Analysis, review, editing

JAY- Analysis

JH- Review, editing

TP- Review, editing, supervision, funding

